# The Italian National Surveillance System for Occupational Injuries: Conceptual Framework and Fatal Outcomes, 2002–2016

**DOI:** 10.1101/2020.06.15.20129726

**Authors:** G Campo, L Cegolon, D De Merich, U Fedeli, M Pellicci, S Pavanello, A Guglielmi, G Mastrangelo

## Abstract

**Background:** A national database of work related injuries was established in Italy since 2002, collecting information on the injured person, his/her work tasks, the workplace as well as risk factors contributing to accident dynamics, according to a model called Infor.Mo.

**Methods:** The present is a qualitative description of occupational fatal injuries, excluding work-related fatal traffic injuries, that occurred in Italy from 2002 to 2016 (15 years).

**Results:** 4,874 victims were registered, all were males, mainly of >51 years of age (50.5%), predominantly self-employed (27.8%) or workers with non-standard contracts (25%). About 18.4% and 17.3% of fatal accidents occurred in micro-enterprises belonging to, respectively, Construction and Agriculture. A wide range of nationalities (59 countries in addition to Italy) was identified. 18.9% work related fatal injuries were due to some form of dangerous energy □ mechanical, thermal, electrical, chemical □ freely present in the workplace. Fall of workers from height (33.5%), heavy bodies falling on workers from height (16.7%) and vehicles exiting their route and overturning (15.9%) were the accidents causing the greatest proportion of occupational fatal injuries. The activity of the injured person made up 43.3% of 9,386 risk factors identified in 4,874 fatalities. Less common risk factors were related to: work equipment (20.2%); work environment (14.9%); the activity of third parties (9.8%); personal protective equipment/clothing (8.0%) and materials (3.7%). The activity of the injured person remained the most relevant contributing factor even when the accident was caused by two or more risk factors.

**Discussion:** Occupational fatal injuries occurred mainly in small size firms. Small companies generally have less resources to catch up with the continuously evolving health and safety at work regulations; moreover, they tend to be less compliant with occupational health and safety regulations since are less likely to be inspected by occupational vigilance services. As a result, the enforcement of regulations to control the occupational risk factors of occupational injuries is seemingly costly and scarcely effective. An alternative approach that is being introduced in Italy relies on the use of economic incentives to promote safe and healthy workplaces.

## INTRODUCTION

### Background

The Italian National Health Service (NHS) aims to ensure heathy and safe working conditions with the involvement and contribution of the employees (law n. 833 of 1978). The Prevention Service for Health & Safety at Work (Italian acronym—SPSAL—the name varies across the various Italian regions) is a section of the Local Health Units (Italian acronym—ASLs) supervising the observation of occupational health and safety (OHS) regulations within the catchment area of the respective ASL. SPSALs, hierarchically subject to the Ministry of Health (MoH) and the Regional Health Services, are the main OHS authorities at the local level in Italy. In compliance with the Legislative Decree 758/1994, SPSAL officers are empowered to act as judicial police officers (Italian acronym—UPG), operative arms of public prosecutors.

In the event of mild occupational non-fatal injuries, the goal of the Italian legislation is primarily ensuring compliance with OHS regulations by supporting business activities. Vigilance actions start with an official workplace inspection of the by SPSAL representatives (technicians and/or occupational doctors), ending up with the issuance of a written report outlining any infringements of OHS norms by the firm. Should the company fail to comply with OHS regulations, SPSAL officers provide technical support, fixing also a deadline to address the workplace violations. A second inspection is then conducted to verify the implementation of the corrections due. If the failure is removed, the infringement is extinguished by payment of an administrative fine. Otherwise, in case of defaulting at the second inspection, the firm is charged with criminal offense, and face legal prosecution as well as coercive measures.

The most severe and fatal occupational injuries are presumed to be criminal offenses. These events cannot be addressed via administrative penalties. Following prompt notification by Accident & Emergency Hospital Services, SPSAL officers immediately conduct investigations in the worksite in order to detect the risk factors that may have contributed or directly cause the accident. Following assessment of the occupational accident, the information collected by UPG is then handed to the prosecutor judge, who will decide whether a prosecution case should be formally opened in court.

The latter decision is influenced by the amount of evidence available; however, the Italian penal system pursues consistent application of the law, allowing limited discretion on judges onto whether to proceed with the case [1,2]. Prosecutors are empowered to request collection of further evidence of regulations breaches where appropriate [3].

The national reference institution for OHS was the Italian National Institute for Prevention and Work Safety (Italian acronym—ISPESL), introduced by law 833/1978 as a body affiliated to the Italian NHS and now incorporated within the National Institute for Insurance against Work Related Accidents (INAIL, Italian acronym). Over 120 years elapsed since its establishment (law n. 80 of 17 March 1898), INAIL has progressively changed its tasks, providing now an integrated system of protection for victims of workplace accidents or occupational diseases, including preventive and research actions at the workplace, medical services, financial support, rehabilitation and reintegration to social life and work. However, the INAIL insurance scheme still compensates workers for occupational injuries and diseases, including payment of workers’ wage. Thus, INAIL is also liable for collecting statistics of work related injuries and occupational diseases.

### Aims

The Italian national surveillance system for occupational fatal injuries was conceptualized and developed within the above framework. The system has been called Infor.Mo (standing for “Infortuni mortali”, work fatalities in Italian). We will herewith describe the development, main features and results achieved by Infor.Mo, through a qualitative analysis of occupational fatal accidents in Italy.

## METHODS

### Development of the system

According to INAIL reports, whilst non-fatal occupational accidents diminished during the nineties, this was not the case for fatal injuries. The collection of further information on causality of fatal accidents was needed for preventive purposes. Therefore, in 2000, in cooperation with the 20 Italian Regions and Autonomous Provinces, ISPESL launched a nation-wide research project to monitor fatal and serious accidents in the workplace. In the same year, INAIL launched a similar project in cooperation with Social Partners, mainly representatives of small and medium industries (Joint Committees).

On 25 July 2002 INAIL, ISPESL and the Conference of Regions and Autonomous Provinces of Italy signed a consensus agreement aimed at integrating the two above projects into a single Integrated Health Information System for Work Prevention, articulated across all 20 Italian Regions. The project was approved and funded by the Italian MoH (MoH Research, Art. 12 e 12 bis D.Lgs. 502/92).

The first complex step of this integrated project was the arrangement of shared working tools. The will of each institution to proceed in partnership and the intense joint activity of their experts produced a unified reference approach. The agreed model is divided in two sections:

- Section1: general description of the events, the injured persons and their workplace/work tasks;
- Section 2: description of the accident dynamics. This section, initially named “Learning by mistakes” (“Sbagliando si impara”, Italian acronym—SSI), was inspired by Northern European experiences [4-6] and developed within the research tasks of ISPESL [7-8].

The model was planned as an information tool to be used during workplace investigations conducted by UPGs to collect evidence on the causes of an accident.

Since the surveillance system Infor.Mo includes several professional figures (UPGs of OHS and researchers of INAIL) and institutions (Regions, ASLs, INAIL), the development of a software application became necessary. An individual central national database was created, archiving all information on major occupational accidents investigated by Infor.Mo. The electronic database is accessible via internet from each local station affiliated to the research network. Data are stored anonymously, adopting various strategies to ensure data protection. An open access website was also created to share information about this project: surveillance; technical materials; publications; training opportunities for companies.

An operational manual was published and distributed to operators/users to instruct about software use and the electronic database. Around 1,000 operators involved in the project were trained in order to achieve or improve their professional competence, maintaining highest standard of homogeneity of skills across different institutions (Regions, ASLs and INAIL).

The INAIL researchers and the operators belonging to Infor.Mo were classified as follows:

- Type A: OHS Officers, whit tasks of accident investigation;
- Type B: Operators with coordination task at regional level;
- Type C: Data managers (INAIL researchers).

Moreover, within companies adopting Infor.Mo, the professional figures appointed to use the model are mainly the Responsible for Prevention and Protection Services and/or employees designated as members of the Protection and Prevention Service.

Each type of operators received different training, following identification of their formative needs, teaching methods, teaching staff, learning assessment tools, evaluation system. There were several updated editions. For SPISAL officers, the training was included in the annual credits for mandatory continuous medical education. In the case of small and medium size industries, the model can find support by trade associations.

Lastly, a Regional Coordination teams (warranting the completeness and quality of information collected at regional level) and a National Coordination team (including representatives of MoH and INAIL) were established. Within the latter, each methodological and organizational aspect of the surveillance system is discussed and agreed.

The initial pilot phase, related to calendar year 2002-2004, was broken down in two parts.

- January 2002 - October 2003, retrospective data collection, from investigations already conducted by SPSAL and local officers of INAIL.
- November 2003 - December 2004: prospective data collection, following the above described surveillance model, with higher standards of data collection, completion and quality.

Data collection has continued until now.

A second objective of Infor.Mo is to support companies in workplace risk assessment, whenever a fatal accident occurs. Furthermore, Infor.Mo has been extended to cover a set of serious non-fatal accidents, with the aim of expanding the detection of risk factors in sectors with frequent, though medium/mild accidents (for example in the manufacturing industry or wood industry). Nonetheless, the Infor.Mo model could be applied to any type of accident, including near misses. The corporate health protection services can use the above model to assess each accident within their companies, highlighting the respective risk factors (to be removed with appropriate interventions) and review the risk assessment.

### Quality checks

The critical points and difficulties encountered by operators/users were mostly related to the application of the Infor.Mo model. Therefore, a further specific professional training was provided in order to support the Regional Scientific Managers and Local Referents of INAIL in carrying out content checks. Moreover, a reference guide was created to define criteria and operational indications to perform the content checks, based on what emerged from this phase of professional update. To further improve the quality of the database, a supervision at the national level was set up for accidents already confirmed at regional level. Numerous meetings were held between operators of all regions to analyze the events and adopt appropriate interpretative solutions. The expected competences of SPSAL officers are updated by annual training. To date, more than 60 courses (10 of which e-learning) have been held, both at national and local level.

### Conceptual framework

The structural components of an injury (all occurring in very short time interval) are:

- the accident;
- the contact (energy exchange); and
- the damage

The reconstruction of the accident dynamics follows the classic backward path used in the judicial investigative process. Starting from the last event in chronological order (the damage), the investigation proceeds to its root causes. Once identified, the three components of the injury (accident, energy exchange, damage) will have to be described by the analyst, who will also categorize them as follows.

The accidents causing work injuries can be classified in two categories.

- Accidents with variation of energy. Episodes of quick and unintended release or transformation or inappropriate application of energy in a work environment, where the contact is the moment of energy exchange between the environment and the worker.
- Accidents with variation of interface energy/worker, where some form of dangerous energy □ namely: mechanical energy (moving gears and machine appliances not adequately isolated); thermal energy (open flames); electrical energy (electrical wires without insulation or with defective insulation); chemical energy (open containers of strongly irritating or caustic substances) □ is normally present in the workplace and directly accessible. These accidents normally occur in workplaces particularly deteriorated and the contact coincides with the injury without prior release of energy.

For both categories of accident, the model Infor.Mo labels the “mode of occurrence” (for example: fall of workers from height, fall of heavy bodies onto workers, vehicles exiting their route and overturning, etc.) according to a list adopted by ESAW (European Statistic of Accidents at Work), a system born in the ‘90s to record work accident data in Europe. For accidents with variation of energy only, Infor.Mo adds the “material agent of an accident”, an information which varies according to the indicated mode of occurrence. For example, being the mode of occurrence “fall of worker from height”, “fall of heavy bodies on workers” or “vehicles exiting their route and overturning”, the material agent describes “from where he fell”, “from where the heavy body fell” or “the vehicle which lost control”, respectively. In summary, the combined information of “mode of occurrence” and “material agent of an accident” provides a specific information on the dynamics of the accident.

The conceptual framework we used to classify factors contributing to a work related injury is a classification scheme allowing a systematic written injury description from the perspective of multiple factors contributing to injuries. This resulted in a matrix consisting of six major categories of contributing factors:

- activity of injured person;
- third party activities;
- machines, tools and plant;
- materials;
- environment;
- personal protective equipment (PPE) and clothing.

Each category can behave as either “determinant” or “modulator” (see below); and each determinant or modulator in turn can be specified as “state” or “process” (see below).

- Determinants and Modulators. Every factor acts as a determinant if it increases the risk of accident. The factor plays as modulator if it is able to worsen the resulting biological damage.
- State and Process. Each factor is defined as “state” if it is present at the beginning of the accident dynamics and remains unchanged during the dynamics. Every factor occurring during the accident dynamic is defined as “process”.
- Safety issue. This variable explains the reasons why a particular category of risk factors was identified by the analyst as an item influencing the accident dynamics under examination. Each of the six categories has its own safety problems, regardless the correspondingrisk factor was either determinant or modulator, state or process.

Numerous sub-categories were developed to classify factors contributing to injury. Overall, 132 sub-categories of contributing factors were identified.

## RESULTS

In the three sections below, we report detailed information on:

- the injured person and workplace/job (table 1);
- the work related injury (tables 2 and 3);
- and the risk factors involved, according to the Infor.Mo model (tables 4 to 9).

Only fatal events passing quality check were selected for the analysis. In an observation period of 15 years (2002 - 2016), the total number of fatal cases and work related accidents was 4,874, an average of 325 cases/year. The number of risk factors detected during workplace investigations was 9,386, an average of 1.9 factors/accident. Data were analyzed descriptively.

**Table 1.** Number (N) and column percentage (%) of fatal injuries in relation to the most important characteristics of the injured worker and their company/occupation.

|  |  |  |  |
| --- | --- | --- | --- |
| FACTORS | CLASSES | N | % |
| Age (years) | 51-65 | 1,477 | 30.7 |
|  | 41-50 | 1,123 | 23.3 |
|  | >65 | 954 | 19.8 |
|  | 31-40 | 600 | 12.5 |
|  | 21-30 | 570 | 11.8 |
|  | Up to 20 | 89 | 1.8 |
|  | Total | 4,813 | 100.0 |
| Citizenship | Italy | 4,091 | 84.6 |
|  | Romania | 208 | 4.3 |
|  | Albania | 208 | 4.3 |
|  | Morocco | 77 | 1.6 |
|  | India | 26 | 0.5 |
|  | North Macedonia | 22 | 0.5 |
|  | Other 54 countries | 279 | 5.8 |
|  | Total | 4,837 | 100.0 |
| Employment Relationship | Permanent (regular) employee | 2230 | 47.0 |
|  | Self-employed without employees | 677 | 14.3 |
|  | Worker in a worker cooperative enterprise | 358 | 7.5 |
|  | Pensioner | 341 | 7.2 |
|  | Irregular and/or precarious worker | 339 | 7.1 |
|  | Self-employed/Owner with employees | 285 | 6.0 |
|  | Non-standard (irregular) employee | 206 | 4.3 |
|  | Fixed-term worker | 163 | 3.4 |
|  | Contributing family worker | 143 | 3.0 |
|  | Total | 4,742 | 100.0 |
| Job | Farmers and Farm Workers | 614 | 14.3 |
|  | Bricklayers, Stonemasons | 501 | 11.6 |
|  | Construction Finishing & Exterior Walls Painting | 292 | 6.8 |
|  | Heavy Goods Vehicle and Truck Drivers | 251 | 5.8 |
|  | Motor Vehicle Mechanics and Repairers | 233 | 5.4 |
|  | Construction of Scaffolding & Tunnel Formworking | 214 | 5.0 |
|  | Electrical Equipment Installers and Repairers | 170 | 4.0 |
|  | Unskilled Construction Workers | 168 | 3.9 |
|  | Carpenters and Joiners in Construction | 157 | 3.6 |
|  | Casters, Welders, Carpentry Fitters | 149 | 3.5 |
|  | Unskilled Farm Workers | 131 | 3.0 |
|  | Other 50 Jobs | 1,422 | 33.1 |
|  | Total | 4,302 | 100.0 |
| Job Seniority | > 3 years | 2,807 | 69.7 |
|  | > 1 year up to 3 years | 459 | 11.4 |
|  | > 1 month up to 6 months | 258 | 6.4 |
|  | Up to 7 days | 206 | 5.1 |
|  | > 6 months up to 1 year | 199 | 4.9 |
|  | 8 days up to 1 month | 100 | 2.5 |
|  | Total | 4,029 | 100.0 |
| Company Size | Up to 9 employees | 2888 | 68.0 |
|  | 16-49 employees | 481 | 11.3 |
|  | 10-15 employees | 410 | 9.6 |
|  | 50-249 employees | 365 | 8.6 |
|  | 250 and more employees | 105 | 2.5 |
|  | Total | 4249 | 100.0 |
| Sectors | Buildings and Civil Engineering Works | 823 | 17.5 |
|  | Crop Farming | 729 | 15.5 |
|  | Building Completion and Finishing | 279 | 5.9 |
|  | Building Installation Services | 237 | 5.0 |
|  | Land, Sea and Air Transport | 209 | 4.4 |
|  | Manufacture of Metal Products | 207 | 4.4 |
|  | Roofing Installation & Roof Construction | 153 | 3.3 |
|  | Agricultural - Related Activities | 148 | 3.1 |
|  | Construction Site Preparation | 132 | 2.8 |
|  | Forestry and Forest Use | 127 | 2.7 |
|  | Other 41 sectors | 1661 | 35.3 |
|  | Total | 4705 | 100.0 |

**Table 2.** Distribution of fatal accidents by mode of occurrence. Number (N); column percentage (%).

| MODE OF ACCIDENT OCCURRENCE (CATEGORIES) | N | % |
| --- | --- | --- |
| Fall of Workers from Height or into Holes | 1635 | 33.5 |
| Fall of Heavy Bodies on Workers | 814 | 16.7 |
| Vehicles exiting their route and overturning | 777 | 15.9 |
| Contact with Moving Vehicles | 360 | 7.4 |
| Unexpected/Improper Starting of Vehicle, Machine, Equipment, etc.. | 297 | 6.1 |
| Contact with Moving Gears | 268 | 5.5 |
| Direct electrical contact | 200 | 4.1 |
| Ejection/emission of solid particles | 155 | 3.2 |
| Development of flames | 92 | 1.9 |
| Other 11 modalities | 276 | 5.7 |
| Total | 4874 | 100.0 |

**Table 3.** Main material agents detected in most frequent modes of occurrence of fatal accidents at work. Number (N); column percentage (%)

|  |  |  |  |
| --- | --- | --- | --- |
| MODE OF OCCURRENCE | MATERIAL AGENT | N | % |
| Fall of Workers from Height or into Holes | Roofs | 483 | 29.5 |
|  | Equipment for Working at Height | 471 | 28.8 |
|  | Other Parts at Height (Balconies, Openings, ...) | 269 | 16.5 |
|  | Lifting and Transport Machines | 64 | 3.9 |
|  | Other Material Agents | 348 | 21.3 |
|  | Total | 1635 | 100.0 |
| Fall of Heavy Bodies on Workers | Lifting and Transport Machines | 142 | 17.4 |
|  | Materials (in Storage Areas) | 127 | 15.6 |
|  | Walls | 80 | 9.8 |
|  | Holes, Excavations, Escarpments | 79 | 9.7 |
|  | Other Material Agents | 386 | 47.5 |
|  | Total | 814 | 100.0 |
| Vehicles exiting their route and overturning | Agricultural and Forestry Machinery | 380 | 48.9 |
|  | Lifting and Transport Machines | 142 | 18.3 |
|  | Earth Moving and Road Works Machines | 102 | 13.1 |
|  | Land vehicles | 92 | 12.5 |
|  | Other Material Agents | 61 | 7.2 |
|  | Total | 777 | 100.0 |

**Table 4.**
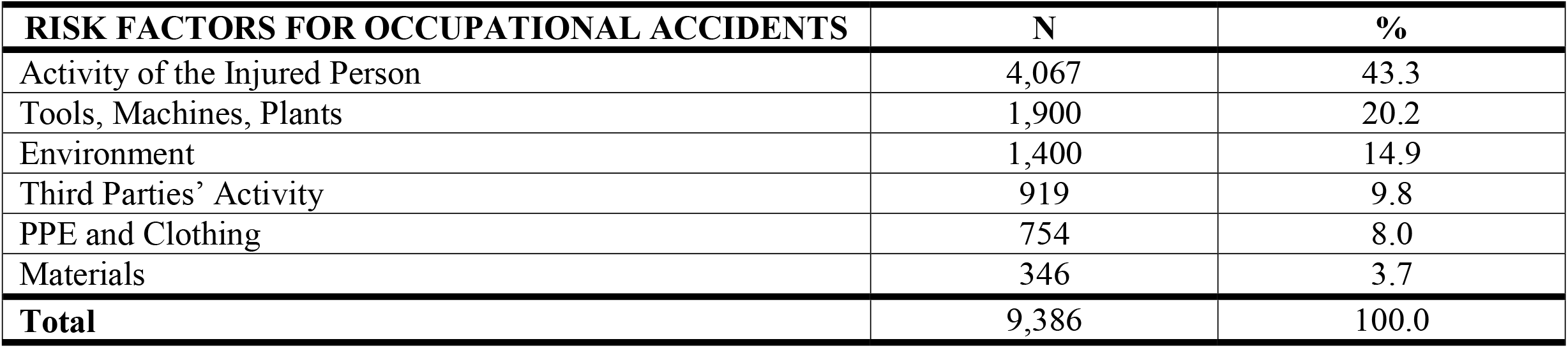
Frequency of risk factors contributing to fatal accident dynamics. Number (N); column percentage (%)

| RISK FACTORS FOR OCCUPATIONAL ACCIDENTS | N | % |
| --- | --- | --- |
| Activity of the Injured Person | 4,067 | 43.3 |
| Tools, Machines, Plants | 1,900 | 20.2 |
| Environment | 1,400 | 14.9 |
| Third Parties’ Activity | 919 | 9.8 |
| PPE and Clothing | 754 | 8.0 |
| Materials | 346 | 3.7 |
| Total | 9,386 | 100.0 |

The number of cases present in the archive Infor.Mo does not cover the total number of fatal work related injuries occurred in Italy. For example, road accidents were excluded because causality investigation was not conducted by SPSAL. On the other hand, some events in the database involved people not formally classified as workers (for example, pensioners or family members) for whom an investigation was conducted because an occupational risk was involved in their injuries.

### Analysis of injured persons

Fatal injuries are predominantly a problem of male workers. According to INAIL statistics, the victims of fatal cases are males in 98% cases. Further main variables traditionally considered in the accident statistics (characteristics of the injured worker, occupation, work task, company sector) are shown in table 1.

The most represented age class of workers was 51-65 years (31%). It is worth mentioning that 12% fatal events still occurred in the most advanced age group (> 65 years), often in the agriculture sector.

Whilst fatal events were much prevalent among Italians (84%), wide variation of nationality was found among foreign workers: 59 different nationalities were identified, predominantly Romanians (4.3%), Albanians (4.3%) and Moroccans (1.6%).

Employees with permanent contract (47%) and self-employed/business owners without employees (14%) were the main working status associated with the accident. At the time of the investigation, 7% injured persons did not have a regular work contract. The proportion of pensioners among occupational fatal injuries is explained by the fact that ASL are liable to perform investigations regardless the person is covered by regular insurance against work related injuries.

Agriculture and Constructions confirmed to be the sectors where occupational accidents produced the greatest damage, with higher rates of fatal accidents, balanced between skilled and generic work tasks. By contrast, the transport industry was under-represented in this database.

Seniority is defined as the length of time an individual has served in a job-task, regardless the number of years of work spent in the company. According to our data, 70% fatal injuries occurred among experienced workers.

68% fatal work related accidents occurred in small companies (<10 employees), normally characterized by lower standards of heath protection.

### Analysis of work accidents

Most fatal injuries are caused by accidents with variation of energy (N = 3953, 81.1% of the total). The accidents with variation of interface energy/worker occurred in less than a fifth (18.9%=921/4874) of total events.

Table 2 shows the distribution of fatal accidents according to mode of occurrence. About half events were caused by workers’ falls from height (33.5%) or heavy bodies falling on workers from height (16.7%). In third position (15.9%) were accidents due to vehicles exiting their lane and overturning. These first three accident modes described about 2/3 of the entire database.

For the first three most frequent modes of occurrence, table 3 shows the corresponding material agents of the accident. Almost a third falls from height (29.5%) occurred from roofs and 28.8% from work equipment at height (primarily scaffolding and portable stairs). Falls of heavy bodies on workers occur mainly from lifting/transporting machines (17.4%) and from storage areas where the materials were not properly stacked (15.6%). Vehicles which lost control were mainly agricultural and forestry machinery (48.9%), lifting/transporting machines (18.3%) and earth moving as well as road works machines (13.1%).

### Analysis of contributing factors

Contributing factors identified in the accident dynamics are analyzed in detail in this section. In all analyses, 9,386 total risk factors were identified for the total 4,874 fatal cases investigated.

Table 4 shows the frequency of risk factors according to the 6 categories proposed by the Infor.Mo model. The activity of the injured person constitutes the most frequently recognized risk factor of fatal accidents (43.3%). Work equipment (20.2%) and environmental related factors (14.9%) represented the following two most frequent factors, whereas the role of third parties’ activity (9.8%), PPE/clothing (8.0%) and materials (3.7%) was more marginal.

Table 5 shows the six categories of contributing factors classified as “determinant” or “modulator”, as well as “state” or “process” according to the role played in the accident dynamics. For these mutually exclusive characteristics, row percentages (rather than column percentages of above tables) were calculated. With the exception of PPE/clothing, which mainly played as modulator (82.5%), all other categories of factors acted largely as “determinants”. The activity of the injured person and third parties’ are by definition measures of process. The other four categories of contributing factors were more frequently classified as “state” rather than “process”. The high frequency of states (especially “tools, machines and equipment”, “materials”, “PPE and clothing”) is of great importance for prevention because the states are by definition identifiable before the accident occurs and completely independent from it. Nevertheless, albeit not always possible and often demanding, processes can still be improved, as endorsed by programs of promotion of quality and good practices.

**Table 5.** Distribution of contributing factors for fatal work accidents according to their role played in the accident dynamics. Number (N); row percentage (%). PPE= Personal protective equipment.

|  |  |  |  |  |  |
| --- | --- | --- | --- | --- | --- |
| FACTORS |  | DETERMINANT | MODULATOR | PROCESS | STATE |
| Activity of the Injured Person | N | 3,380 | 687 | 4,067 | 0 |
|  | % | 83.1 | 16.9 | 100.0 | 0.0 |
| Third Parties' Activity | N | 878 | 41 | 919 | 0 |
|  | % | 95.5 | 4.5 | 100.0 | 0.0 |
| Tools, Machines, Plants | N | 1471 | 429 | 274 | 1626 |
|  | % | 77.4 | 22.6 | 14.4 | 85.6 |
| Materials | N | 332 | 14 | 110 | 236 |
|  | % | 96.0 | 4.0 | 31.8 | 68.2 |
| Environment | N | 1269 | 131 | 176 | 1224 |
|  | % | 90.6 | 9.4 | 12.6 | 87.4 |
| PPE and Clothing | N | 132 | 622 | 117 | 637 |
|  | % | 17.5 | 82.5 | 15.5 | 84.5 |
| Total | N | 7462 | 1,924 | 5,663 | 3,723 |
|  | % | 79.5 | 20.5 | 60.3 | 39.7 |

Table 6 shows the distribution of the safety problems identified for each of the six categories of contributing factors. The security problems associated with “Activity of injured persons” include procedure or operative errors (75.7%), incorrect (18.5%) or improper (5.8%) use of equipment. Records which did not present any safety issue (N=44) were excluded. Each type of security issue could be broken down into detailed operational aspects. For example, 38.8% procedural errors were due to incorrect practices usually adopted within the company and 30.9% to the lack of OHS courses in the workplaces. Unfortunately, this evidence is weakened by a percentage of missing values as high as 43% (= (1741□3046)/3046, see figures in bold, table 6). Similar considerations apply to the security problems related to “Third-parties’ activities”, where the percentages of missing observations for procedural errors, incorrect use of equipment and improper use of equipment were 43%, 49% and 56% respectively (14 records with missing information on security problems were excluded). Nonetheless, these results indicate the need to improve training of workers.

**Table 6.**
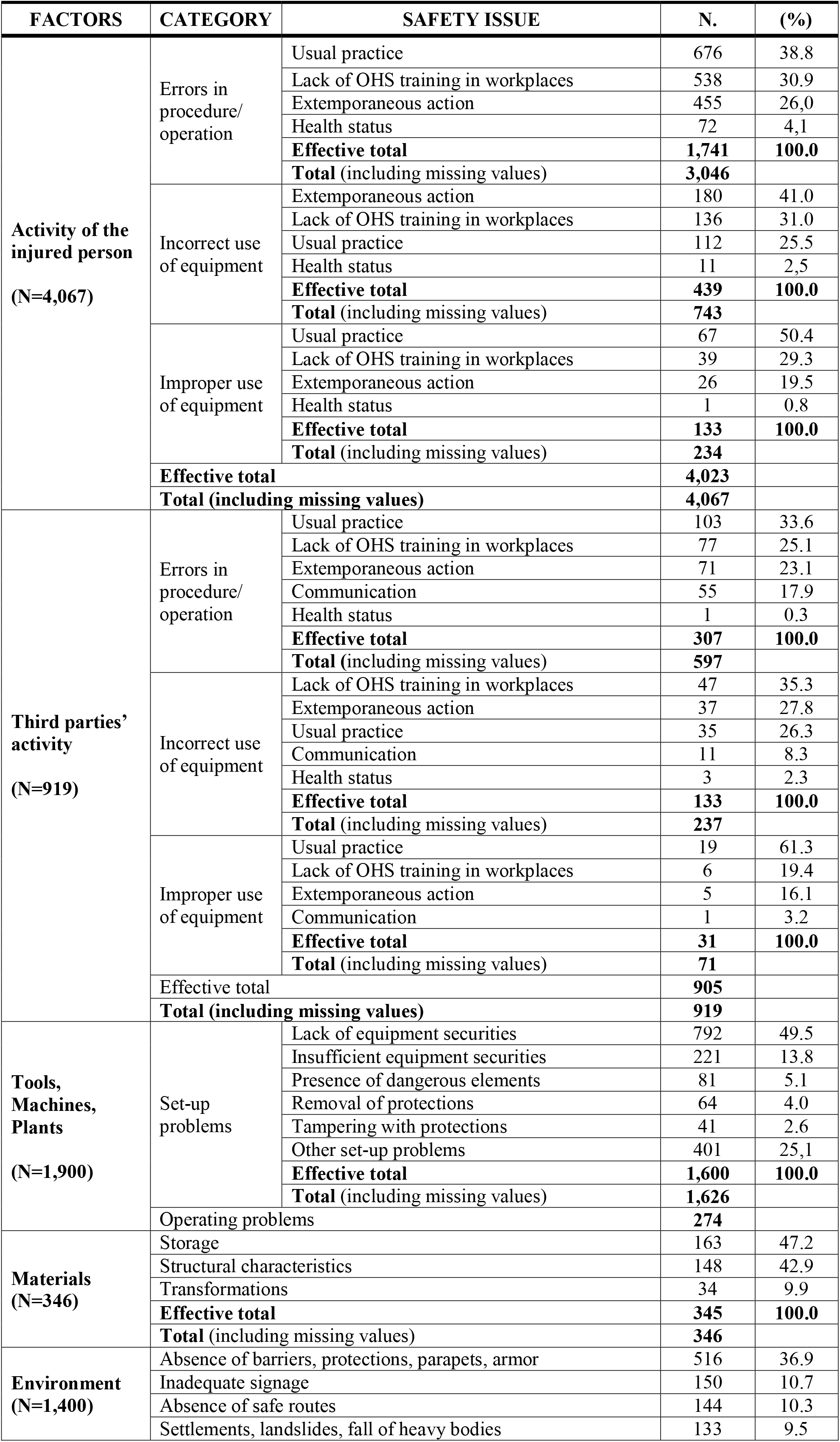
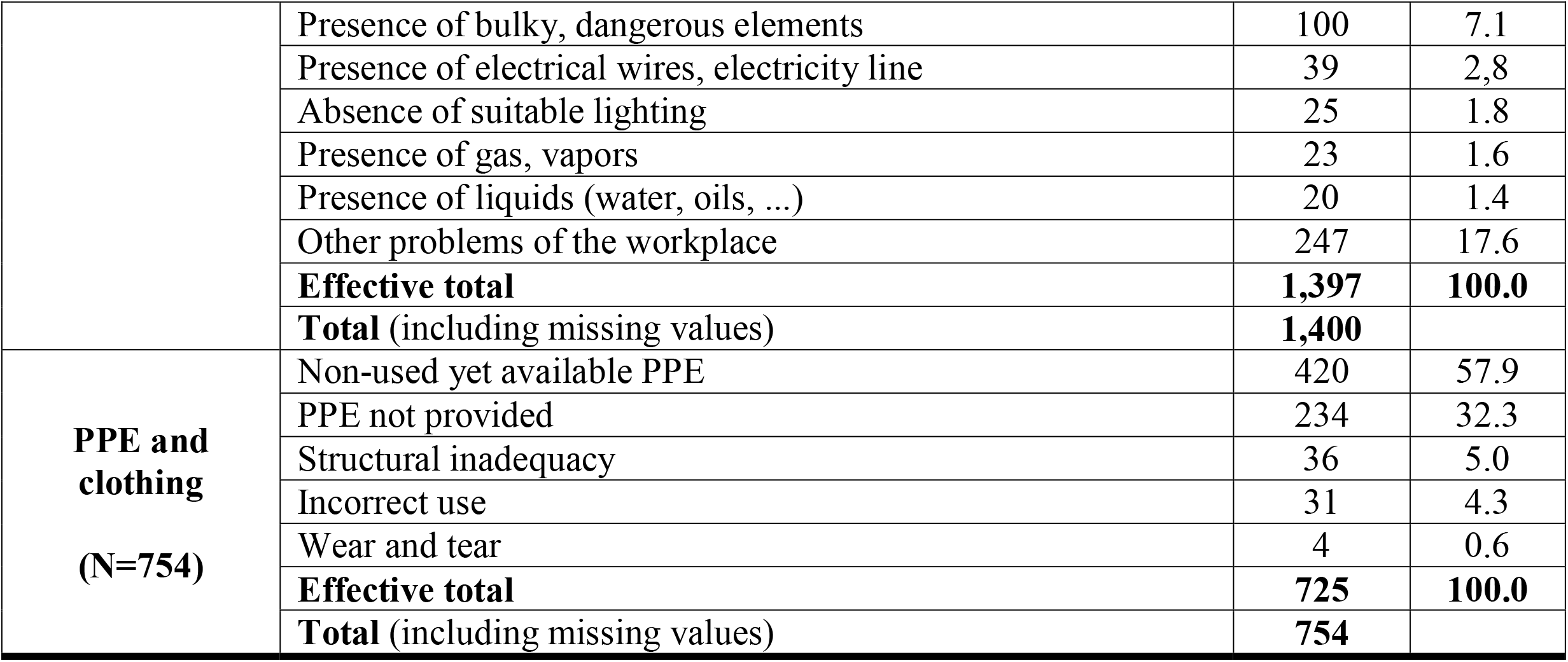
Factors contributing to fatal work accidents by category and safety issues. Number (N); column percentage (%).

As for “Tools, machines and plants”, the set-up problems are much more frequent (85.6%) than the operating problems (14.4%). In detail, 72% set-up problems are due to lack, inadequacy, removal or tampering of protections necessary to work in safe conditions. Interestingly, out of 1,900 observations only 26 are missing values. These results are in line with the preponderance of the states with respect to the processes reported in table 5 and confirmed the high predictability and preventability of the accidents examined. The contributing factor “Materials” presented three safety problems (1 missing), related to their storage (47.2%), their structural characteristics (42.9%) or their transformations (9.9%). The safety problems related to the “Environment” are mainly the absence of barriers, protections, parapets, armor (36.9%), along with inadequate signals and absence of safe routes (21%); the latter issue affects the movements of pedestrians and vehicles. Three missing values were found among the 1,400 observations on workplace conditions. Concerning “PPE and clothing”, there are two safety issues (29 missing): non-use but available PPE (57.9%); and PPE not provided (32.3%).

Table 7 shows a two-way classification of fatal work accidents, stratifying the 6 categories of risk factors by their number causing the accident. The last column of table 7 displays the distribution of accidents by number of risk factors (column percentage). If only one factor was identified, the corresponding category of contributing factors was the activity of the injured person (in 58.4% fatal accidents). The activity of the injured person was the most relevant contributing risk factor even in the case the accident was caused by two or more risk factors. Overall, 4,067 out of 9,386 risk factors (43.3%) concerned activity of the injured person.

**Table 7.** Two-way classification of fatal occupational accidents by number of factors originating the accidents (rows) and category of contributing factors (columns). Number (N), percentage (%). PPE= Personal protective equipment.

|  |  |  |  |  |  |  |  |  |  |
| --- | --- | --- | --- | --- | --- | --- | --- | --- | --- |
| N. of Risk Factors |  | CATEGORIES OF CONTRIBUTING FACTORS |  |  |  |  |  |  | accidents<br>(column %) |
|  |  | Activity<br>of injured<br>person | Third<br>parties'<br>activity | Tools,<br>machines,<br>plants | Materials | Environm<br>ent | PPE and<br>clothing | All<br>categories |  |
| 1 | N | 938 | 140 | 252 | 51 | 206 | 19 | 1,606 | 33.0 |
|  | (row %) | 58.4 | 8.7 | 15.7 | 3.2 | 12.8 | 1.2 | 100.0 |  |
| 2 | N | 2,081 | 389 | 931 | 129 | 633 | 367 | 4,530 | 46.5 |
|  | (row %) | 45.9 | 8.6 | 20.6 | 2.8 | 14.0 | 8.1 | 100.0 |  |
| 3 | N | 863 | 305 | 512 | 89 | 417 | 268 | 2,454 | 16.8 |
|  | (row %) | 35.2 | 12.4 | 20.9 | 3.6 | 17.0 | 10.9 | 100.0 |  |
| 4 | N | 151 | 66 | 128 | 33 | 104 | 74 | 556 | 2.9 |
|  | (row %) | 27.2 | 11.9 | 23.0 | 5.9 | 18.7 | 13.3 | 100.0 |  |
| 5 | N | 33 | 18 | 47 | 23 | 38 | 26 | 185 | 0.8 |
|  | (row %) | 17.8 | 9.7 | 25.4 | 12.4 | 20.5 | 14.1 | 100.0 |  |
| 6 | N | 1 | 1 | 2 | 0 | 2 | 0 | 6 | 0.0 |
|  | (row %) | 16.7 | 16.7 | 33.3 | 0.0 | 33.3 | 0.0 | 100.0 |  |
| 7 | N | 0 | 0 | 28 | 21 | 0 | 0 | 49 | 0.1 |
|  | (row %) | 0.0 | 0.0 | 57.1 | 42.9 | 0.0 | 0.0 | 100.0 |  |
| Total | N | 4,067 | 919 | 1,900 | 346 | 1,400 | 754 | 9,386 | 100.0 |

The 9,386 risk factors detected in 4,874 work related accidents equalled 1.9 factors/accident. Therefore, from the collection of 9,386 contributing factors, we selected all combination of two factors. The total number of 2-combinations, f(2), is equal to summary of the binomial coefficients

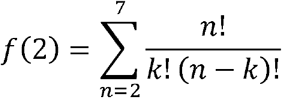

where k = 2 (pairs of factors) and n = 2 to 7 (number of risk factors by fatal event; there were 7 fatalities with 7 risk factors).

Table 8 reports the frequencies of combinations of two risk factors of fatal accidents. The association “Acvitity of the injured person” & “Tools, Machines, Plants” was the most common, collecting 22.4% of all 6,085 associations. “Activity of the injured person” & “Environment” came next, making up 14.5% fatalities. It is worth noting that according to Infor.Mo model each fatality could have 2 or more risk factors of the same category (e.g. two factors both classified as “Activity of the injured person”), which may represent different entities to be treated separately from a preventative perspective.

**Table 8.** **Frequencies of combination of 2 risk factors of occupational fatal injuries. Number (N), percentage (%). PPE= Personal protective equipment**.

|  |  |  |  |  |  |  |  |  |
| --- | --- | --- | --- | --- | --- | --- | --- | --- |
| RISK FACTORS |  | Activity of the injured person | Activity of third parties | Tools, Machines, Plants | Materials | Environment | PPE and clothing | Total N (column %) |
| Activity of the injured person | N | 405 (6.7) | 572 (9.4) | 1,362 (22.4) | 209 (3.4) | 882 (14.5) | 562 (9.2) | 3.992 (65.6) |
|  | (% of total) |  |  |  |  |  |  |  |
| Activity of third parties | N |  | 66 (1.1) | 223 (3.7) | 52 (0.9) | 223 (3.7) | 72 (1.2) | 636 (10.5) |
|  | (% of total) |  |  |  |  |  |  |  |
| Tools, Machines, Plants | N |  |  | 236 (3.9) | 159 (2.6) | 253 (4.2) | 236 (3.9) | 884 (14.5) |
|  | (% of total) |  |  |  |  |  |  |  |
| Materials | N |  |  |  | 32 (0.5) | 77 (1.3) | 63 (1.0) | 172 (2.8) |
|  | (% of total) |  |  |  |  |  |  |  |
| Environment | N |  |  |  |  | 130 (2.1) | 246 (4.0) | 376 (6.2) |
|  | (% of total) |  |  |  |  |  |  |  |
| PPE and clothing | N |  |  |  |  |  | 25 (0.4) | 25 (0.4) |
|  | (% of total) |  |  |  |  |  |  |  |

Based on data of table 8, table 9 displays the combinations of each risk factor with another risk factor in decreasing order of frequency. As can be seen, the activity of the injured person is of utmost importance for fatal occupational accidents because it is always present in each combination of two risk factors.

**Table 9.** **Most frequent associations between 2 risk factors of fatal occupational accidents, by decreasing frequency. PPE= Personal protective equipment**.

|  |  |
| --- | --- |
| RISK FACTORS | MOST FREQUENT ASSOCIATION FOR FATAL EVENTS |
| Activity of the injured person | Tools, Machines, Plants |
| Activity of third parties | Activity y of the injured person |
| Tools, Machines, Plants | Activity of the injured person |
| Materials | Activity y of the injured person |
| Environment | Activity of the injured person |
| PPE e clothing | Activity of the injured person |

## DISCUSSION

### Key results

There was no model in Italy that could guarantee the uniformity of analysis for occupational accidents. A single model □ one of the most interesting aspects of the project □ was difficult to define because it needed to overcome different established practices and experiences of the operators/inspectors. Nontheless, Infor.Mo model has been generally believed or recognized to be valid or correct compared to the previous methods. Therefore, a national database of occupational injuries was established in Italy since 2002, collecting information on injured persons and workplace/work tasks as well as risk factors contributing to accident dynamics, according to the model Infor.Mo. The model is currently in use by SPSAL officers in their investigations on behalf of the prosecutor judge. Having passed accurate quality controls, the present national database of 4,874 occupational fatal injuries constitutes a solid base for their description and interpretation of fatal accidents, excluding traffic work-related injuries, occurring in Italy from 2002 to 2016 (15 years).

Victims involved are male workers, mainly of >51 years of age (50.5%), which were predominantly self-employed (27.8% being micro-entrepreneurs without/with employees or member of worker-cooperative) or non-standard workers (25% summing up pensioner, contributing family worker and irregular or precarious/fixed term worker). About 18.4% and 17.3% of fatal accidents occurred in micro-enterprises belonging to, respectively, Construction and Agriculture, sectors already known for the high risk of injuries. A wide range of nationalities (59 countries excluding Italy) was identified among injured workers.

Less than a fifth (18.9%) accidents were due to some form of dangerous energy □ mechanical, thermal, electrical, chemical □ freely present in the workplace; the relevant more common causes were contact with moving gears (5.5%) and direct electrical contact (4.1%). Since a dangerous energy, capable of causing serious damage or death, was directly detectable and manageable during the risk assessment, the latter accidents were easily predictable and preventable. On the other hand, about 40% of accidents were due to indicators of “state”, yet present at the beginning and remaining unchanged during the accident dynamics. Tools, machines, plants and, moreover, environment acted mainly as “state” risk factor. The assumption that occupational accidents are the result of chance would frustrate any attempt to prevent them. Rather, occupational injuries are preventable because they are events generally determined by pre-existing worksite conditions [9].

Workers’ falls from height (33.5%), heavy bodies falling on workers from height (16.7%) and vehicles exiting their route and overturning (15.9%) were about 2/3 of the entire database. Agricultural and forestry machinery were about half of the vehicles that lost control; others were lifting/transporting machines, earth moving as well as road works machines.

Activity of the injured person accounted for 43.3% of 9,386 risk factors detected in 4,874 fatal injury cases. Less common risk factors were related to: work equipment (20.2%); environment (14.9%); third parties’ activity (9.8%); PPE/clothing (8.0%) and materials (3.7%). The activity of the injured person remained the most relevant contributing factor even when the accident was caused by two or more risk factors. The combinations of two distinct risk factors always comprised the activity of the injured person. Concerning the safety problems associated with activity of injured persons, errors in procedure/operation, incorrect use of equipment and improper use of equipment ranked first (75.7%), second (18.5%) and third (5.8%), respectively.

### Limitations

#### Lack of information on personal risk factors

The risk of occupational injuries increases with work-related factors along with personal factors, such as age and sex, smoking [10-16], physical inactivity [17-18], alcohol consumption [10-11], and obesity [11,16], as well as prior health-related conditions such as cardiovascular disease [17-21], mental illness [12,19,21-22], musculoskeletal conditions [11,19-20], diabetes [19-21, 23-24], and asthma [19,21]. Human factors also include workers’ action/behavior related to safety, skills, communication and available supervision [25-26], inter-personal and hierarchical relationships, corporate context, unions, internal and external work competition, etc. [26]. Since human beings are not perfect, accidents sustained by workers’ errors – either by poor attention, negligence or recklessness – call for further training and behavioral changes in terms of increased attention during job tasks [27].

The role of personal factors could not be confirmed in the present study because, apart from age and sex, the database do not contain further information on worker’s characteristics. Nonetheless, activity of the injured person was the most important risk factors detected in 4,874 fatal injury cases. An important number of missing data, however, did not allow to recognize whether the cause underlining the activity of the injured person was of occupational origin (“usual practice”, “lack of OHS training in workplaces”) or a personal risk factor (“extemporaneous action” or “health status”). The same applies to third parties’ activity. Probably, accident analysts (UPGs of SPSAL) focused attention on law violation or safety issue. Under this assumption, priority was given to quest for responsibilities, an approach mainly oriented to repressive aspects rather than prevention.

#### Lack of denominators

The ratio between events recorded in a population sample (numerator) and subjects in the same sample (denominator) with simultaneous assessment of time at risk permits to compare the risk of work accidents among quantitatively different samples of population. In Italy, for insured workers, there is currently no information available on the composition by gender, age, ethnic origin, contractual typology and other potentially interesting characteristics. For this reason we have used a simple descriptive analysis. These findings therefore need further confirmation in studies with precise denominator information.

### Interpretations of findings

In the present investigationy, fatal injuries occurred mainly in small size firms with deteriorated workplaces and use of irregular work, suggesting that some employers seek out offsetting reductions in their labor costs. In agreement with present findings, higher rates of fatal/severe accidents in smaller worksites than larger ones were observed among the 17,481 occupational fatalities investigated in the US by the Occupational Safety and Health Administration during 1992-2001 [28]. Although this seems to suggest that smaller workplaces generate higher occupational risks, several additional explanatory factors may be involved [28]. First, since they cannot benefit from large scale economies, small companies have less resources to catch up with the continuously evolving Health & Safety norms [28-29]. Second, the burden of safety and occupational regulations is heavier on small business activities than large ones [28]. Third, smaller worksite are frequently outreach and less likely to be inspected by occupational vigilance services [28].

To assess the effects of occupational safety and health regulation and legislation enforcement activities, such as inspections, for preventing work related injuries and occupational diseases, a review included 23 studies: two randomised controlled trials on 1,414 workplaces, two controlled before-after studies on 9,903 worksites, one interrupted time series with 6 outcome measurements, 12 panel studies and 6 qualitative studies with 310 participants. Although with low level of evidence, workplace inspections were seemingly capable to reduce accidents more in the long run than in the short run. The effect size was stronger with focused ather than general inspections. The effect of fines and penalties remained undefined. Since the quality of the evidence was low to very low, these conclusions were tentative and could be easily amended in the future by improved studies [30].

There was an urgent need for better designed evaluations to establish the effects of existing and novel enforcement methods. A more recent systematic review of literature covering the years 1966□2017, including 45 journal articles and 16 reports of the gray literature, was undertaken to assess whether OHS legislative and regulatory policies could reduce industrial injuries and fatalities [31]. The review indicates that legislative and regulatory policy may reduce injuries and fatalities and improve compliance with OHS regulation. The evidence for improvement was moderately strong [31].

Nevertheless, concerning intervention studies on small enterprises, a review of studies published before 2006 reported a lack of evaluation, both in terms of effect and practical applicability. The most effective preventive approaches seemed to be simple and low cost solutions, disseminated through personal contact [32]. A later systematic review was undertaken to identify effective occupational health and safety interventions for small businesses. Five studies of medium or high quality were identified. A combination of training and safety audits, and a combination of engineering, training, safety audits, and a motivational component, showed a limited evidence in improving safety outcomes. However, stronger levels of evidence are required to draw appropriate recommendations [33].

Workers’ falls from height, heavy bodies falling on workers from height, and vehicles exiting their route and overturning were the accidents causing the greatest proportion of fatal injuries either in the present study (from 2002 to 2016) and in the initial pilot phase, related to years 2002-2004 [34]. Their distribution was similar in the two periods, being 33.5% versus 31.9%, 16.7% versus 15.1%, and 15.9% versus 12.7% for, respectively, fall of workers from height, objects falling on workers and loss of control of vehicles. Most of the above accidents occurred in small firms belonging to Construction and Agriculture.

Despite being a critical business for all countries in the world, constructions are featured by high injury risk. According to van der Molen et al [35], the vast majority of technical and human factors and organisational interventions recommended by standard safety textbooks, consultants and safety courses, have not been adequately evaluated. To assess the effectiveness of interventions for preventing work related injuries among construction workers, a systematic review was conducted using published findings from the earliest available dates through June 2006 [36]. Findings from a safety-campaign study and a drug-free-workplace study indicated that both interventions significantly reduced the level and the trend of injuries. Three studies evaluating legislation did not found decreased levels of non-fatal or fatal injuries in the construction industry [36].

In a more recent systematic review on 17 interventional studies (14 interrupted time series and 3 before-after studies) collected until 1 April 2017, an immediate effect was measured in the year immediately after the intervention and a sustained effect was assessed as change in time trend before and after the intervention. The association between regulatory measures at national and local level and immediate as well as sustained effect on fatal and non-fatal occupational accidents was also not consistent and safety training did not translate into significant reduction of non-fatal injuries. Economic incentives to companies may reduce the rates of non-fatal injuries from falls, and multifaced interventions may significantly decrease the initial and sustained accident rate at company level, albeit not at regional scale. Whilst a multifaced drug-free workplace intervention at company level might decrease non-fatal injuries in the year following the implementation and to smaller extent also in the subsequent years, the implementation of occupational health services did not reduce fatal and non-fatal accidents [37].

Beside construction industry, agriculture was the other main area of concern for occupational fatalities in the present study. Likewise, among 335,000 yearly fatal occupational injuries worldwide, 170,000 regarded agricultural workers [38]. In a systematic review on interventional studies in agriculture until June 2006, 3 randomized controlled trials on 4,670 adult participants and 2 RCT on 6,895 children participants did not show any significant reduction if the rates of occupational accidents. Whilst educational interventions may not significantly decrease agriculture injuries, monetary incentives may be effective in reducing them. Regulations enforcing the introduction of protective devices on all tractors in Sweden were not beneficial in reducing fatal injuries, while the same measures for new tractor machines translated instead into less fatal accidents [39-40].

### Perspectives

The latter finding suggests that engineering controls (e.g. a new tractor) have higher impact than minor changes in work practices (e.g. adopting rollover protective gear applied on an old tractor) in reducing the risk of occupational injuries. Neverthless, several employers seem more prone to contain labour costs than improving technology to enhance productivity. Fatal injuries occurred mainly in small size firms with deteriorated workplaces and employment of undocumented workers. Since they cannot benefit from large scale economies, small companies have less resources to catch up with the continuously evolving health and safety regulations. An approach recently introduced in Italy relies upon economic incentives granted by INAIL to companies through formal calls for application for projects and interventions aimed to promote safe and healthy workplaces [41]. It is still unclear whether financial incentives are more successful than OHS regulations to ensure healthy worksitesand new evidence is needed. Randomization of firms in two groups, intervention companies and controls (“waiting list” companies), could allow an unbiased evaluation of financial incentives in preventing occupational injuries; but if OHS regulations are not adequately implemented, as often occurs for small companies, they are unlikely to be effective [42].

## Data Availability

The dataset analysed in the current study is not publicly available, but may be obtained from the corresponding author on reasonable request.

## Author Contributions

Conceptualization and design, G.C. and G.M.; Analysis, G.C., A.G., D.D. and M.P; Writing, G.M., L.C. and S.P.; Supervision, G.M., U.F., A.G. and L.C.

## Acknowledgments

We wish to thank Dr Luciano Marchiori (Department of Prevention, Aulss 9, Veneto Region, Verona, Italy) for his preliminary work on data that persuaded authors to undertake the present work.

